# The Role of Adverse and Positive Childhood Experiences on Mood Challenges in Youth

**DOI:** 10.1101/2025.05.09.25327239

**Authors:** Hyeeun K. Shin, Scott C. Leon, Olga Vsevolozhskaya, Xiaoran Tong, John S. Lyons

## Abstract

Accumulative adverse childhood experiences (ACEs) have been shown to increase the risk of physical and mental health issues in children. More recently, accumulative positive childhood experiences (PCEs) have been shown to lessen the negative impact of ACEs; however, the research to date has rarely included children in foster care. We investigated the cumulative association between ACEs and PCEs among youth in foster care (*N=19,888*) ages 5-18 in a midwestern state between 2011-2023 on the likelihood of mood challenges (MC), including anger control, anxiety, depression, and affective dysregulation. We further stratified by age, sex, and race. ACEs were associated with a greater likelihood of MCs, while PCEs were associated with decreased likelihood of these outcomes. The offset effect of PCEs was greater for affect dysregulation and anger control compared to anxiety and depression, highlighting the potentially distinct role of positive experiences with adults on children’s emotion regulation.

## Introduction

Adverse Childhood Experiences (ACEs) are traumatic events that occur in childhood and include abuse, neglect, and other trauma-inducing occurrences (e.g., natural disasters). Due to their prevalence and impact, the Centers for Disease Control and Prevention (CDC) considers ACEs to be a top public health concern (CDC, 2023). Approximately one in six adults has experienced four or more ACEs, with four or more ACEs serving as the classical cut-point at and above which rates of negative outcomes rise significantly (Felitti et al., 1998). Negative outcomes among adults sampled retrospectively reporting ACEs include severe mental health and behavioral issues such as depression, suicidal ideation/attempts, anxiety, smoking, and substance misuse (Austin, 2018; Hughes et al., 2017). These challenges can, in turn, lead to chronic health conditions in adulthood, such as coronary heart disease, stroke, and cancer (Merrick et al., 2019; Petruccelli et al., 2019).

Early research on ACEs and their sequelae primarily focused on the adult population; however, subsequent studies have demonstrated that ACEs can have deleterious effects long before adulthood. For example, Elmore and Crouch (2020) analyzed data from the National Survey of Children’s Health – a nationally representative, 50-state survey of children and adolescents, -- and their findings revealed that children aged 8-17 with two or more ACEs demonstrated significantly higher odds of current anxiety (Odds Ratio (OR):1.8; 95% Confidence Interval (CI):1.5-2.2) and depressed mood (OR:2.6; 95% CI:2.0-3.3) compared to those with fewer than two ACEs. Overall, existing research strongly indicates that the impact of ACEs is both significant and widespread, with adverse effects emerging in childhood and persisting into adulthood (Bielas et al., 2016; Lyons et al., 2023). Given international research indicating that one in seven youth worldwide experiences a mental health issue, representing 15% of the global disease burden in this age group (WHO, 2024), understanding protective factors that mitigate the negative impact of ACEs on mental health must remain a public health priority.

Fortunately, not all children exposed to ACEs develop negative long-term outcomes in adulthood. Resilience in these cases has been strongly linked to Positive Childhood Experiences (PCEs). PCEs -- including positive relational experiences with the nuclear family, extended family, peers, school, and other supportive people in the community -- are associated with well-being, reduced risky behavior, and higher social-emotional functioning in adulthood (Bethell et al., 2019; Daines et al., 2021). For example, Bethell and colleagues (2019) found that among adults with ≥4 ACEs, approximately 60% of those reporting 0-2 PCEs had current depression or poor mental health. In contrast, only 20% of those with ≥4 ACEs but 6-7 PCEs experienced depression or poor mental health in adulthood. Research suggests PCEs demonstrate particularly strong protective effects against depression relative to ACEs (Han et al., 2023). Beyond depression, studies document PCEs’ promotive and protective benefits across multiple domains in ACE-exposed populations, including substance use disorders and body image (e.g., (Crandall et al., 2020)), attachment (e.g., (Anderson, 2021)), emotion regulation (e.g., (Hanson et al., 2024)).

Historically, studies examining the relationship between ACEs, PCEs and mental health relied predominantly on adult retrospective report on childhood experiences alongside assessments of current mental health status or functioning. However, the past five years have seen a dramatic increase in studies examining the relationship between ACEs, PCEs, and mental health outcomes in childhood and adolescence (e.g., (Bunting et al., 2023; Elmore & Crouch, 2020; Qu et al., 2022; Samji et al., 2024; Scholtes & Cederbaum, 2024; Wang et al., 2021)). Although this remains a relatively new area of inquiry, an international picture is beginning to emerge suggesting that PCEs demonstrate comparable or greater predictive power than ACEs for mental health outcomes in childhood and adolescence, albeit operating in opposite directions. Accumulating evidence from diverse cultural contexts -- including Northern Ireland (Bunting et al., 2023), China (Qu et al., 2022), Canada (Samji et al., 2024) and the United States (Elmore & Crouch, 2020; Scholtes & Cederbaum, 2024; Wang et al., 2021) -- consistently demonstrates the promotive and protective benefits of PCEs against cumulative ACE exposure. These findings persist across various study designs ranging from nationally representative (Elmore & Crouch, 2020; Hinojosa & Hinojosa, 2023) to at-risk population studies (Wang et al., 2021).

Despite these advances, significant gaps remain in the literature. While adult studies have consistently discover a wide breadth of promotive and protective benefits for PCEs that counterbalance the deleterious effects of ACEs, child and adolescent research has primarily focused on internalizing disorders, with depression being most extensively studied (Bunting et al., 2023; Elmore & Crouch, 2020; Qu et al., 2022; Samji et al., 2024; Wang et al., 2021), followed by anxiety (Bunting et al., 2023; Qu et al., 2022; Wang et al., 2021). Beyond depression and anxiety, emerging evidence demonstrates associations between ACEs/PCEs in relation to suicidal ideation and self-harm (Bunting et al., 2023); well-being and life satisfaction (Samji et al., 2024); psychological distress (Scholtes & Cederbaum, 2024); and disruptive behavior (Hinojosa & Hinojosa, 2023). Notably absent from this growing literature are clinical phenomena frequently observed in youth, such as emotional dysregulation and poor anger control, despite their prevalence in child and adolescent populations. Existing evidence suggests that strong positive relationships with family and school, along with proper parental supervision, serve as some of the most effective protective factors for antisocial and illegal behavior (Cleveland et al., 2010), fostering practical anger management skills (Brezina, 2010). These findings suggest that PCEs might be effective in counter-acting or buffering the association between ACEs and emotional dysregulation and anger control difficulties (Almeida et al., 2024; Feiler et al., 2023) shown to exist in the literature.

Another gap in the existing ACEs/PCEs and mental health literature involves the lack of research with the child welfare population. This oversight is particularly significant given that children in foster care likely have higher levels of ACEs and may have unique resilience pathways compared to children in the general population. While existing research suggests that resilience for children in the general population is tied more to consistent parenting and stable community relationships, foster youth often experience more fragmented social support systems (e.g., (Nuñez et al., 2022)). This difference suggests that variability in PCEs may be a particularly powerful predictor of mental health outcomes among children in foster care. Moreover, the existing literature uses an operationalization of ACEs consistent with the types of adversity more commonly seen in the general population, such as divorce, witnessing violence, and the presence of a substance-abusing parent. While physical abuse is frequently included in these measures, other clinically significant ACEs (like neglect, emotional abuse, and sexual abuse) are less commonly assessed. Therefore, the current literature, using samples with far fewer and less severe ACEs than what is experienced by youth with child welfare involvement, may be less generalizable to children in foster care.

Additionally, a picture is still developing in the literature among children and adolescents regarding whether PCEs function primarily as “promotive” factors in relation to ACEs (beneficial regardless of ACE exposure) or “protective” factors (buffering against ACE-related risk). For example, Bunting et al. (2023) found that ACEs and PCEs acted significantly but independently (i.e., with no interaction effect between the two) in predicting depression, anxiety, self-harm, and suicidal ideation, making PCEs “promotive.” PCEs, therefore, were associated with better mental health outcomes regardless of ACE exposure and did not appear to act as a buffer to increasing ACEs. However, contrasting evidence from Elmore et al. (2020) demonstrated a dual protective role of PCEs: they were not only associated with lower odds of depression as a main effect but also buffered the impact of ACEs on depression. Samji and colleagues (2024) further nuanced this picture, with PCEs showing protective effects for depression (i.e., significant ACEXPCE interaction) and promotive effects for anxiety, well-being, and life satisfaction (i.e., main effects only). This pattern of differential effects was similarly observed by Qu and colleagues (2022), who found a significant buffering effect of PCEs on depression among high-risk adolescents (those with four or more ACEs) but no comparable buffering effect against anxiety. These collective findings suggest that more research is needed to better understand the ways and contexts in which PCEs impact mental health and resilience in the presence of ACEs.

### Current Study

This study examines the differential associations between cumulative ACEs and PCEs in relation to four mood challenges -- depression, anxiety, anger control, and emotional dysregulation – analyzed both individually in separate models and in an overall model with a composite of all four. Unlike previous research, we examine these associations using ACEs that are more inclusive of the experiences of children in the foster care system. Our investigation pursues three primary aims: (1) examine the association between ACEs and mental health outcomes, (2) examine the association between PCEs and mental health outcomes, and (3) examine the role of PCEs as a potential moderator of the association between ACEs and mental health outcomes. We further disaggregate our findings by age (5-12 and 13-18), race (Black and White), and sex (female and male) to identify potential subgroup variations.

## Materials & Methods

The source population comprises youths served by the child welfare agency (CWS) of a large Mid-Western state between February 2010 and December 2022. The primary data source is the Child Adolescent Needs and Strengths (CANS) 2.0 assessment (Lyons, 2009), which is required by state regulations to be administrated to youth under state custody every six months. The final study sample passing quality control included 19,888 children aged 5–18 years, with balanced representation across younger (50.4% aged 5–12) and older (49.6% aged 13–18) cohorts. The sample was predominantly male (53.3% male, 46.7% female) and of White race (63.4% White, 36.6% Black). The study employed a per-person random cross-sectional design (Archary et al., 2015) to ensure representativeness of the population at various stages of engagement with the CWS over the entire period of 12 years (2010-2022) for which data were available.

### Child and Adolescent Needs and Strengths (CANS)

The CANS, built upon the conceptual framework of Transformational Collaborative Outcomes Management (TCOM), is a person-centered, evidence-based tool used in 50 states and internationally to measure the clinical and functional status of children and adolescents (Lyons, 2009; Lyons, 2022). The version of CANS used presently has 150 indicators rated on four-point action levels from ‘0’ to ‘3’ across eight core domains: trauma experience, adjustment to trauma, life functioning, school, child and family cultural factors, child behavioral/emotional needs, child risk behaviors, and child strengths.

The four action levels represent an increasing action priority. For items measuring needs, 0 = No evidence, no need for action; 1 = Watchful waiting/prevention, history, suspicion, or contention; 2 = Action, the need is interfering with functioning; and 3 = Immediate/Intensive Action, the need is dangerous or disabling. For items measuring strengths, 0 = Centerpiece strength (the strength is present); 1 = Useful strength (the strength is present); 2 = Identified strength but must be built to be useful; and 3 = No strength has been identified.

Prior research has demonstrated that the CANS contains reliable indicators (Dilley, 2007; Rachel L. Anderson, 2003). Specifically, Anderson et al. (2003) report an inter-rater reliability score of 0.85 for the CANS total scale. We re-coded the rating to a binary format to enhance our statistical analysis and reporting robustness. So, ratings of “0” or “1” are categorized as “0 = non-actionable,” indicating that no immediate action is required, while ratings of “2” or”3” are classified as”1 = actionable,” signifying that action is necessary. To ease the interpretation, we reverse coded indicators of strengths, from “1=actionable” to “0 = strength undeveloped”, and from “0=non-actionable” to “1=strength present”.

### Mood Challenges (MCs) and Total Mood Challenge (TMC)

The outcomes of interest are mood challenges identified as the actionable/non-actionable statuses of four indicators in the child behavioral/emotional needs domain of the CANS: affect dysregulation, anger control, anxiety, and depression (Supplementary Table 1). These 4 outcomes are emotional reactions that can often impair daily life (Weinstein & Mermelstein, 2007) and are represented in the CANS assessment as irritable mood, mood change, and mood swings. The Cronbach’s alpha of 4 mood challenges gave a score of 0.70, indicating acceptable internal consistency. Additionally, we created a fifth outcome named Total Mood Challenge (TMC) ranged 0 to 4, as the sum of actionable status of the 4 indicators.

### Adverse Childhood Experiences (ACEs)

ACEs are a key predictor. In the study, ACEs is a score in 0 to 9, defined cumulatively as the sum of actionable status of 9 indicators in the trauma experience domain of the CANS, namely, sexual abuse, physical abuse, neglect, emotional abuse, medical trauma, natural or manmade disaster, witness to family violence, witness to community activity, and witness to criminal activity (Supplementary Table 1). While our ACE indicators differ slightly from those defined in Felitti’s (1998) original work, which formed the foundation for many ACE studies, the CANS allows for the capture of a broader range of adverse life experiences (Shin et al., 2024).

### Positive Childhood Experiences (PCEs)

PCEs are another key predictor we focus on. This score aggregates actionable indicators, ranging from 0 to 9, within the child strengths domain of the CANS. Specifically, we selected 9 out of 17 items related to youths’ external environment, relationship with others, and interaction with the community as our PCE indicators. The nine indicators of PCEs include relationship permanence, family-nuclear, family extended, positive peer relations, educational setting, recreational activities, community life, spiritual/religious, and natural supports (Supplementary Table 1). The Cronbach’s Alpha of the 9 indicators was 0.77, suggesting acceptable internal consistency among indicators of PCEs.

### Covariates

In this study, we only included age, sex, and race as covariates either as regression terms or stratification criteria to avoid overfitting and to enhance the generalizability of results (Rochefort-Maranda, 2016). Age is grouped into 0 = “school-aged children (5-12 years old)” and 1 = “teenagers (13-18 years old)”; sex is categorized as 0 = “female” and 1 = “male.”; race is categorized into two primary groups: 0 = “Black” and 1 = “White” since other racial groups that constitute only 8% of the source population were excluded from the study population. Each covariate coded as 0 will be treated as the baseline reference.

### Analysis

We provide descriptive statistics of the baseline characteristics of our sample population, including chi-Square tests for categorical variables (i.e., age, sex, and racial group) and ANOVA for continuous variables (i.e., ACEs and PCEs) to assess the demographical differences between individuals with and without mood challenges.

We used linear regression to model the main and interaction effects of PCEs and ACEs on the total mood challenge (TMC) outcome while controlling for age, sex, and race. The model is formulated as:

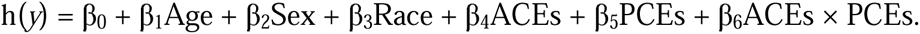

On the right-hand side, the core independent variables were counts of ACEs and PCEs ranging from 0 to 9 while ACEs × PCEs being product of the two; Age, Sex, and Race are binary variables defined under “Covariates.” On the left-hand side, the dependent variable *y* is the outcome of Total Mood Challenge (TMC) ranging from 0 to 4 - putting through the link function *h,* which, for TMC, is the trivial identity function. The regression coefficients were estimated by R/4.1.2 Statistical Software (Supplementary Table 2), among which, β_4_ and β_5_ depict the main effects per additional ACE and PCE on the outcome independent of one another; β_6_ depicts the interaction, a.k.a., mutual effect modification between ACEs and PCEs and vice versa. In particular, a negative β_5_ indicates the promotive benefit of gaining PCEs; a negative β_6_ indicates the protective benefit of gaining PCEs in terms of mitigating the impact of ACEs, and conversely, a positive β_6_ indicates the diminishing benefit of gaining PCEs in the presence of ACEs, while (β_5_ + β_6_ ACEs) depicts the overall effect of gaining one additional PCE in the presence of ACEs. The coefficients allowed us to estimate the expected changes in TMC given any number of PCEs and ACEs; a 10 by 10 heatmap (Figure 1, Supplementary Figure 1) was then created to visualize the expected changes in mean TMC given PCEs and ACEs ranged from 0 to 9. Each tile’s color gradient - progressing from dark blue (indicating TMC reduction) through neutral white (no change) to dark red (TMC increase) - illustrated the estimated mean TMC changes associated with each combination of PCE and ACE counts. The tiled heatmap offers two key analytical advantages: (1) the relative areas of blue versus red immediately reveal whether PCEs collectively outweigh ACEs in their impact on TMC, and (2) the pattern of the democratization line (where the impact of PCEs and ACEs exactly cancel out each other) demonstrates whether additional PCEs confer protective of diminishing benefit.

**Figure 1.**
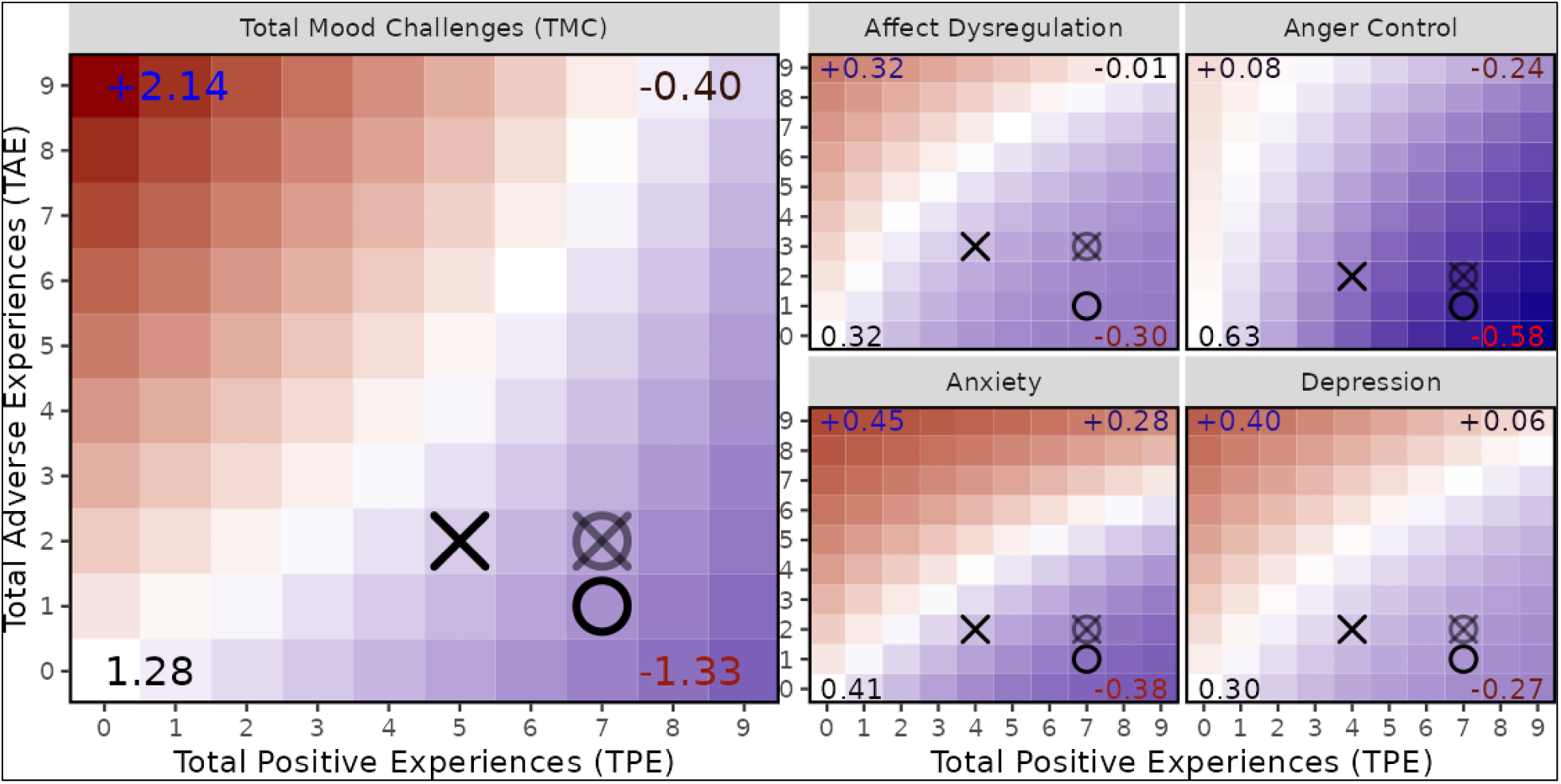
Cumulative Effect of Positive vs Adverse Childhood Experiences on Mood Challenges - by Total Sample. The **x/y-axis** represent exposures of 0 to 9 PCEs and ACEs, respectively; **(0, 0)** depicts the estimated outcome of a baseline individual (i.e., age 0-5, male, and white) with 0 PCE and 0 ACE; **cooler/warmer color at (x, y)** depicts, relative to the baseline, the estimated effect of x-PCEs and y-ACEs combined, to either mitigate or escalate the said outcome; **(0, 9)** at the top-left shows the effect of 9 ACEs without PCE, likewise, **(9, 0)** at the bottom-right show the effect of 9 PCEs without ACE. The **democratization line** denotes mutually neutralizing numbers of PCEs and ACEs; **cool area being larger or smaller than warm area** suggests PCEs overcomes ACEs in general (e.g., Anger Control) or vice versa (e.g., Anxiety); **left-yielding democratization** implies protective benefit of PCEs in the presence of increasing number of ACEs (e.g., Total Mood Challenges); **bottom-yielding democratization** implies diminishing benefit of additional PCEs (e.g., Affect Dysregulation and Anxiety); **straight democratization** implies the lack of mutual moderation between PCEs and ACEs (e.g., Depression). The "⨯" and "⭘" mark medium exposures to PCE/ACE among cases and non-cases, respectively (for Total Mood Challenges, "cases" are individuals with at least one out of four mood challenges); "⦻" denotes a counterfactual case whose PCEs were built up to the median number among non-cases, despite the inability to remove ACEs.

Next, treating each of the four mood challenges (MCs) as a binary outcome, we used logistic regression to model the main and interaction effects of PCEs and ACEs on MC, while controlling for age, sex, and race. The structure of the model formula was unchanged, but the dependent variable *y* was the prevalence of an MC put through the logit link function *h*(*y*) = log [*y* / (1-*y*)] (converts the prevalence to logged odds). The exponent of β_5_ depicts the odds ratio of an MC among youth having one additional PCE versus not, while holding ACE at zero; the exponent of (β_5_ + β_6_ ACEs) depicts the overall odds ratio of an MC among youths having one additional PCE versus not in the presence of ACEs. Again, a negative β_5_ indicates promotive benefit of gaining PCEs, a negative β_6_ indicates protective benefit of gaining PCEs in mitigating the impact of ACEs, and a positive β_6_ indicates diminishing benefit of gaining PCEs in the presence of ACEs. Likewise, the regression coefficients (Supplementary Table 2) allowed us to estimate and visualize changes in the prevalence of each MC across the full range of PCEs and ACEs (Figure 1, Supplementary Figure 1).

Finally, we stratified the samples by age (5-12 or 13-18), sex (Male or Female), and race (White or Black) to explore the varying impacts of PCEs and MCs across demographic groups. Again, we used logistic regression to model the main and interaction effects of PCEs and ACEs on the logged odds of each MC but controlling every other two demographic factors when one was chosen to stratify the samples. The interpretation of regression coefficients (Supplement Table 2) and the estimation and visualization of expected changes in the prevalence of each MC across the range of ACEs and PCEs (Figure 2, Supplementary Figure 2) were similar to those in the previous full-sample analysis.

**Figure 2.**
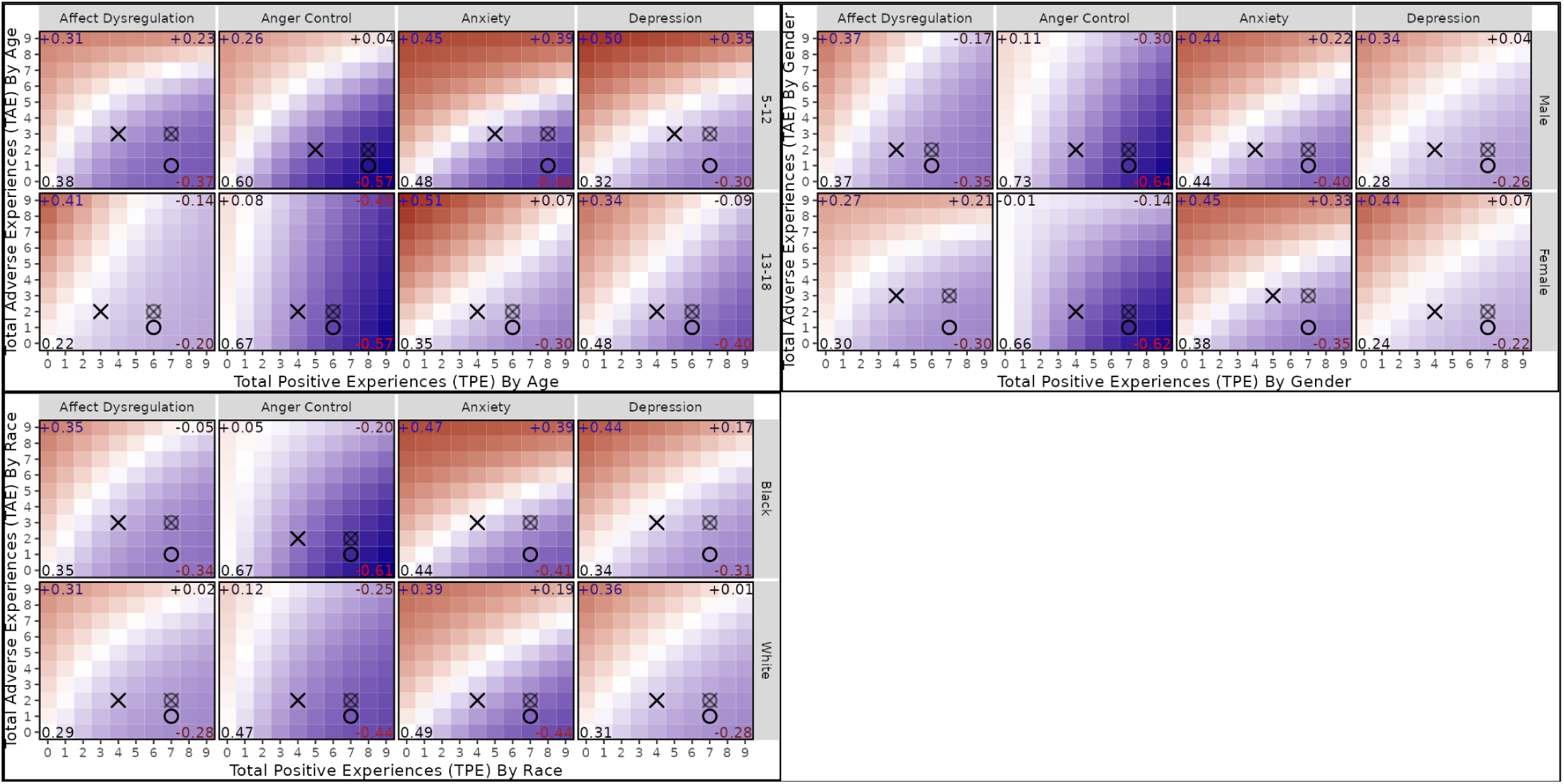
Cumulative Effect of Positive vs Adverse Childhood Experiences on Mood Challenges – Stratified by Age, Gender, and Race. The 3 blocks show effects stratified by **Age (Top-left)**, **Gender (Top-right)**, and **Race (Bottom-left)**. The **x/y-axis** represent exposures of 0 to 9 PCEs and ACEs, respectively; **(0, 0)** depicts the baseline (i.e., age 0-5, male, and white) with 0 PCE and 0 ACE; **cooler/warmer color at (x, y)** depicts, relative to the baseline, estimated effect of x-PCEs and y-ACEs combined, to either mitigate or escalate the outcome; **(0, 9)** at the top-left shows the effect of 9 ACEs without PCE, likewise, **(9, 0)** at the bottom-right show the effect of 9 PCEs without ACE. The democratization line denotes mutually neutralizing numbers of PCEs and ACEs; cool area being larger or smaller than warm area suggests PCEs overcomes ACEs in general (e.g., Anger Control) or vice versa (e.g., Anxiety); left-yielding democratization implies protective benefit of PCEs in the presence of ACEs (e.g., Affect Dysregulation among 13-18); bottom-yielding democratization implies diminishing gain of additional PCEs (e.g., Affect Dysregulation among 0-5); straight democratization implies mutually unaffecting PCEs and ACEs (e.g., Depression among female). The “⨯” and “⭘” mark medium exposures to PCE/ACE among cases and controls, respectively; “⦻” denotes a counterfactual case whose PCEs were built up to the median number among controls, despite the inability to remove ACEs

To ensure the reliability of findings gained from the main analysis, we conducted sensitivity analysis by re-drawing four more random cross-sectional data sets and repeating all the above modeling. This resulted in estimates consistent with the main (Supplementary Table 2 for the results of both main and sensitivity analysis).

## Results

### Population Characteristics

We present the sample characteristics in Table 1. Among 19,888 youths, anger control was the most prevalent mood challenge (MC) with a case rate of 24.8%, followed by anxiety (23.6%), depression (18.8%), and affect dysregulation (9.8%). Age was significantly associated with MCs, with adolescents (13-18 years) demonstrating higher rates than children (5-12 years; P < 0.001). Sex differences emerged in specific MCs: males were overrepresented in anger control (P < 0.001) and affect dysregulation (P < 0.001) cases, while females predominated in depression cases (P < 0.001). Racial disparities were also evident, with Black youths showing higher rates of anger control relative to White youths (P < 0.001), whereas White youths had higher rates of anxiety (P < 0.002) and depression (P < 0.002).

**Table 1.**
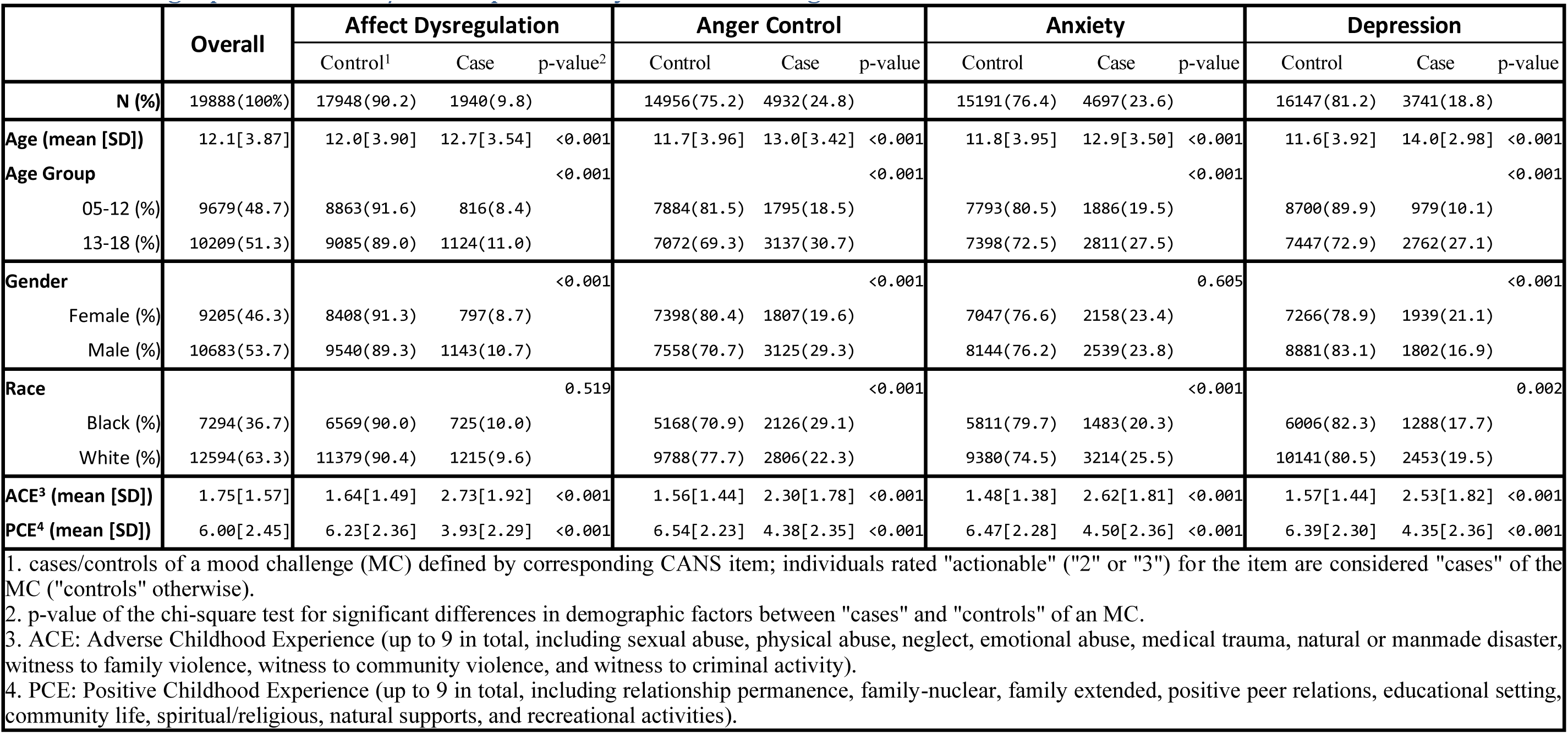
Demographics and PCE/ACE exposures by mood challenges.

|  | Overall | Affect Dysregulation |  |  | Anger Control |  |  | Anxiety |  |  | Depression |  |  |
| --- | --- | --- | --- | --- | --- | --- | --- | --- | --- | --- | --- | --- | --- |
|  |  | Control<sup>1</sup> | Case | p-value<sup>2</sup> | Control | Case | p-value | Control | Case | p-value | Control | Case | p-value |
| **N (%)** | 19888(100%) | 17948(90.2) | 1940(9.8) |  | 14956(75.2) | 4932(24.8) |  | 15191(76.4) | 4697(23.6) |  | 16147(81.2) | 3741(18.8) |  |
| **Age (mean [SD])** | 12.1[3.87] | 12.0[3.90] | 12.7[3.54] | <0.001 | 11.7[3.96] | 13.0[3.42] | <0.001 | 11.8[3.95] | 12.9[3.50] | <0.001 | 11.6[3.92] | 14.0[2.98] | <0.001 |
| **Age Group** |  |  |  | <0.001 |  |  | <0.001 |  |  | <0.001 |  |  | <0.001 |
| 05-12 (%) | 9679(48.7) | 8863(91.6) | 816(8.4) |  | 7884(81.5) | 1795(18.5) |  | 7793(80.5) | 1886(19.5) |  | 8700(89.9) | 979(10.1) |  |
| 13-18 (%) | 10209(51.3) | 9085(89.0) | 1124(11.0) |  | 7072(69.3) | 3137(30.7) |  | 7398(72.5) | 2811(27.5) |  | 7447(72.9) | 2762(27.1) |  |
| **Gender** |  |  |  | <0.001 |  |  | <0.001 |  |  | 0.605 |  |  | <0.001 |
| Female (%) | 9205(46.3) | 8408(91.3) | 797(8.7) |  | 7398(80.4) | 1807(19.6) |  | 7047(76.6) | 2158(23.4) |  | 7266(78.9) | 1939(21.1) |  |
| Male (%) | 10683(53.7) | 9540(89.3) | 1143(10.7) |  | 7558(70.7) | 3125(29.3) |  | 8144(76.2) | 2539(23.8) |  | 8881(83.1) | 1802(16.9) |  |
| **Race** |  |  |  | 0.519 |  |  | <0.001 |  |  | <0.001 |  |  | 0.002 |
| Black (%) | 7294(36.7) | 6569(90.0) | 725(10.0) |  | 5168(70.9) | 2126(29.1) |  | 5811(79.7) | 1483(20.3) |  | 6006(82.3) | 1288(17.7) |  |
| White (%) | 12594(63.3) | 11379(90.4) | 1215(9.6) |  | 9788(77.7) | 2806(22.3) |  | 9380(74.5) | 3214(25.5) |  | 10141(80.5) | 2453(19.5) |  |
| **ACE<sup>3</sup> (mean [SD])** | 1.75[1.57] | 1.64[1.49] | 2.73[1.92] | <0.001 | 1.56[1.44] | 2.30[1.78] | <0.001 | 1.48[1.38] | 2.62[1.81] | <0.001 | 1.57[1.44] | 2.53[1.82] | <0.001 |
| **PCE<sup>4</sup> (mean [SD])** | 6.00[2.45] | 6.23[2.36] | 3.93[2.29] | <0.001 | 6.54[2.23] | 4.38[2.35] | <0.001 | 6.47[2.28] | 4.50[2.36] | <0.001 | 6.39[2.30] | 4.35[2.36] | <0.001 |

1. cases/controls of a mood challenge (MC) defined by corresponding CANS item; individuals rated "actionable" ("2" or "3") for the item are considered "cases" of the MC ("controls" otherwise).
2. p-value of the chi-square test for significant differences in demographic factors between "cases" and "controls" of an MC.
3. ACE: Adverse Childhood Experience (up to 9 in total, including sexual abuse, physical abuse, neglect, emotional abuse, medical trauma, natural or manmade disaster, witness to family violence, witness to community violence, and witness to criminal activity).
4. PCE: Positive Childhood Experience (up to 9 in total, including relationship permanence, family-nuclear, family extended, positive peer relations, educational setting, community life, spiritual/religious, natural supports, and recreational activities).

When breaking down PCEs and ACEs indicators (Supplementary Figure 3), we observed that, except for neglect and witness to family violence, adolescents (age 13-18) had greater exposure to both ACEs and lack of PCEs than school-aged children (age 5-12). No significant difference emerged in exposure to PCEs/ACEs by sex and race (figures not included).

### Effect of PCE and ACE Combined

Figure 1 (and Supplementary Figure 1) depicts the effect of PCEs and ACEs on MCs estimated by full-sample models adjusted for age, sex, and race. PCEs alone showed statistically significant promotive effects on all 5 outcomes (β_5_ = −0.148 for Total Mood Challenge, −0.405 for affect dysregulation, −0.390 for anger control, −0.334 for anxiety, and −0.303 for depression, all P < 0.001). ACEs alone showed significant opposite effect on 4 out of 5 outcomes except anger control (β_4_= 0.237 for Total Mood Challenge, 0.150 for affect dysregulation, 0.243 for anxiety, and 0.186 for depression, all P < 0.001; for anger control, β_4_ = 0.041, P = 0.094), the heatmap showed that the negative effect of 9 ACEs could be offset by as few as 3 PCEs. When considered together, an average PCE was found to overcome the impact of an average ACE on Total Mood Challenge (TMC), particularly with protective benefit among individuals with a higher number of ACEs (β_6_ = −0.015, P < 0.001). However, for anxiety and depression, the effect of an average PCE was surpassed by an average ACE, and PCEs showed signs of diminishing effectiveness (β_6_ = 0.024 for anxiety, 0.016 for depression, both P < 0.001) in the presence of high numbers of ACEs.

### Experiences on Mood Challenges - Stratified by Age, Gender, and Race

Figure 2 (and Supplementary Figure 2) depicts the impact of PCEs and ACEs on MCs as estimated by regression models with samples stratified by age, sex, and race. The overall influence of PCEs and ACEs was consistent with previous models based on the entire sample. However, differences were observed based on age: the effects of PCEs on the 4 MCs were universally more promotive and less diminishing among teenagers (age 13-18) than their younger counterparts (age 5-12). A sex difference was also observed regarding affect dysregulation, where the effect of ACEs was more dominant and unyielding among females than males. For race, the effect of ACEs on affect dysregulation was slightly more pronounced among White individuals than for Black individuals.

## Discussion

This study examined how adverse (ACEs) and positive childhood experiences (PCEs) jointly influence mood challenges in foster care youth. Analyses revealed that PCEs’ cumulative protective effect outweighed ACEs’ detrimental impact on total mood challenges (TMC); for example, a child with 9 PCEs and 9 ACEs was predicted to have fewer TMC relative to baseline. However, upon examining mood indicators separately (i.e., affect dysregulation; anger control, anxiety; and depression), we observed variability in the relative influence of adverse and positive childhood experiences regarding their associations with specific mood challenges. Specifically, PCEs more effectively offset ACEs’ detrimental effects for affect dysregulation and anger control than they did for anxiety and depression. Further, significant ACEs × PCEs interactions emerged across most models, indicating that the compensatory benefits of PCEs are stronger at lower levels of ACEs and attenuated at higher levels of ACEs.

### ACEs and PCEs: Affect Dysregulation and Anger Control versus Depression and Anxiety

The results presented in the heat maps (Figures 1-2) and the beta coefficients for ACEs and PCEs (Supplementary Table 3a) across the four mood challenges -- affect dysregulation, anger control, anxiety, and depression -- suggest that, overall, PCEs are stronger predictors than ACEs for affect dysregulation and anger control. Specifically, in the affect dysregulation model, the beta coefficient for PCEs is approximately three times larger than that for ACEs. Similarly, in the anger control model, the PCEs coefficient is about ten times higher than the ACEs coefficient. Although PCEs also showed higher coefficients than ACEs in the depression and anxiety models, the differences were substantially smaller compared to those observed in the affect dysregulation and anger control models.

Challenges associated with affect dysregulation and anger control have been characterized as reactive and under-controlled emotion responses, while anxiety and depression have been characterized as containing over-learned and internalized emotion regulation strategies (e.g., rumination and avoidance; (Cole et al., 2019)). Our findings broadly indicate that PCEs may play a relatively more significant role in helping children and adolescents regulate reactive emotion experiences and may be relatively less impactful in helping youth alter maladaptive internalizing mood disorders that have over-controlled, intrapersonal components to their etiology and maintenance, such as depression and anxiety. However, it is important to note that PCEs played a compensatory role across all four mood challenge models; the differences we discuss here are relative, not absolute.

Affect dysregulation and anger control difficulties often lead to interpersonal difficulties but can also be modified by positive interpersonal experiences. Previous research highlights the beneficial impact of stable, nurturing, and calm interactions with adults on children who experience difficulties with anger and affect regulation. For example, positive interpersonal experiences with adults -- particularly those who demonstrate the ability to tolerate intense emotional displays and label the emotion experience through reflection -- can teach children to better regulate their own emotional experiences, thereby leading to its modification in future settings. This process has been documented across multiple published work. For example, in the *mentalization* literature, positive experiences with adults have the potential to reflect the child’s intense emotional experience back to the child, but less intensely, thereby showing that the emotion is safe and capable of regulation (Fonagy & Target, 2003).

Parallel to the emotion regulation literature, the intervention literature emphasizes *interpersonal* approaches for clinical presentations related to anger control and affect dysregulation and emphasizes *intrapersonal* approaches for depression and anxiety. For example, parent management strategies (e.g., Parent-Child Interaction Therapy; (McNeil et al., 2010)) focus on teaching parents how to prevent emotion dysregulation through reflection and praise and to respond to disruptive emotional and behavioral displays through the calm and regulated application of negative punishment (e.g., time outs) strategies. Alternatively, treatments for anxiety and depression focus on the intrapersonal experience of negative automatic thoughts, core beliefs, rumination, and avoidance behaviors (i.e., Cognitive Behavioral Therapy; e.g., (James et al., 2020)).

### ACEs and PCEs: Interaction Effects

The protective effects of PCEs appear to interact with the level of ACEs. Specifically, PCEs appear to be beneficial in protecting against mood challenges at lower levels of ACEs but begin to wane in significance as ACEs increase. This type of interaction between a putative risk and putative protective factor has been termed by Luthar and colleagues (2000) as *Protective-Reactive*, which occurs when a protective factor lessens the impact of a risk factor but does not eliminate its impact. Protective-reactive effects are common in research measuring the accumulation of ACEs. One protective variable that has been studied extensively and is noteworthy for demonstrating a protective-reactive pattern among mood disorders is social support (e.g., (Rueger et al., 2016)), which is relevant to our study given that our PCEs refer to positive relationship experiences. Our results appear to fit within a broader context in the existing literature, demonstrating that while social support is critical, it might not be enough on its own to be protective in the context of extreme levels of adversity. At higher levels of adversity, other factors, such as intrapersonal protective factors (e.g., self-esteem, coping, optimism), may also be needed to protect against the experience of mood (e.g., (Buchanan et al., 2023)), particularly anxiety and depression.

### Differences Across Age, Sex, and Race

Teenagers (age 13-18) showed greater exposure to both ACEs and PCEs compared to school-aged children (age 5-12; Supplementary Figure 3). However, school-aged children exhibited a higher risk of experiencing the negative effects of ACEs on their mood challenges compared to teenagers (Figure 2, Supplementary Figure 2). This observation contrasts with some research showing that teenagers are more likely than school-aged children to experience cumulative and higher effects of ACEs (Meeker et al., 2021). However, our findings align with other research indicating that there are greater well-being differences between samples of younger children with complex trauma versus children with neglect compared to older children with complex trauma versus neglect (Huguenel et al., 2021). School-aged children are still developing their language and emotional regulation skills, which might make them more susceptible to mood challenges stemming from the immediate effects of ACEs due to their developmental stage (Reilly & Downer, 2019), while teenagers typically possess more advanced cognitive abilities that enable them to process and cope with adversity. Over time, they often become more independent, which strengthens their resilience, including seeking support outside the family and engaging in self-directed behaviors to help regulate their emotions (Silvers et al., 2012). Furthermore, both school-aged children and teenagers are likely to have externalizing behavior, such as physical symptoms or significant changes in mood challenges (Petersen, 2024); however, teenagers may be more skilled at concealing their emotional challenges and tend to internalize their feelings (De France & Hollenstein, 2019).

Comparing females and males, there was little variation in how ACEs and PCEs were associated with anxiety and depression. Regarding affect dysregulation, PCEs were highly beneficial for both females and males; however, females were more susceptible to the effects of ACEs. The anger control results revealed an opposite pattern. While, once again, both males and females benefited from PCEs, males were more negatively influenced by ACEs. Our findings are consistent with previous research, which suggests that females are more vulnerable to emotional dysregulation in response to adversity, while males are more likely to exhibit externalized behaviors, such as anger and impulsivity, when exposed to childhood adversity (Leban & Delacruz, 2023; Van Wyk, 2023). It is important to note that boys and girls often experience different types and patterns of ACEs. A study using latent class analysis to examine the co-occurrence of ACEs in females and males has revealed that the types of ACEs experienced may differ by sex. This study indicates that females often face more complex and varied experiences, frequently encountering multiple types of ACEs at the same time, compared to males (Haahr-Pedersen et al., 2020). These observed sex differences in responses to risk factors, such as ACEs, may be influenced by sociobiological interaction mechanisms, including female sex hormones, which are closely linked to issues like emotional dysregulation (Whitaker et al., 2021).

Regarding race, there were no differences in how ACEs and PCEs impacted anger control. For affect dysregulation, while PCEs were beneficial for both Black and White children and adolescents, the White youth were vulnerable to the impacts of ACEs. In terms of anxiety and depression, PCEs provided weaker positive effects for both Black and White youth; however, the Black youth were more affected by ACEs. Aldao et.al (2010) found that maladaptive emotional regulation (e.g., affect dysregulation), which is related to a broad range of mental health issues, has been consistently implicated in anxiety and depression (Sloan et al., 2017). Both Black and White individuals experience mental health challenges due to ACEs; however, Black individuals often are exposed to a higher number of ACEs, which may lead to sustained stress (Zhang & Monnat, 2022). Nevertheless, Black individuals have been shown to be more likely to rely on community support and social networks (Jacob et al., 2023). Despite this, the cumulative impact of ACEs can be more detrimental to anxiety and depression for Black individuals due to additional stress stemming from systemic inequity (Wilhoite et al., 2024). While White individuals may be more likely to have immediate responses to ACEs, Black individuals often face ongoing stress due to the cumulative impact of both ACEs and sustained oppression at a cultural level.

### Limitations

While the large population-based sample is a strength, this study has a number of limitations. First, our study includes a child welfare sample from one state, potentially limiting generalizability. Second, although adverse and positive childhood experiences can lead to mood challenges, they are certainly not the only factors, and many of them were not measured in this study. For example, biological factors also play a significant role in the development of mood challenges, including genetic, epigenetic, gene by environment interaction, and hormonal variables (Bhangle et al., 2013; Juruena et al., 2020). Adding to the complexity, these variables interact with age (e.g., pre-versus post-pubertal) and sex. Third, we measured the cumulative impact of ACEs and PCEs; therefore, we could not determine the varying effects of each specific indicator of ACE and PCE. For example, prior work has found that sexual abuse is a far more powerful negative predictor of mental health than other ACE items (Briggs et al., 2021). This study provided a broad and nomothetic view of the relationship between ACEs and PCEs, which, despite its advantages, has the weakness of not offering a view into the nuanced and complex relationship between children and their environments across development. Future work should seek to provide an idiographic, person-centered perspective on some of the specific ACEs and PCEs studied here, holding other ACEs and PCEs constant.

Fourth, our dataset includes the period of the COVID-19 pandemic, which has the potential to increase or decrease mood challenges (Matsumoto et al., 2023) and adverse (Racine et al., 2024) and positive childhood experiences (Crouch et al., 2024) among young individuals. During the COVID-19 pandemic, many children and adolescents had fewer opportunities to experience PCEs, which could have impacted the variability of our PCEs variable, with associated impacts on our outcomes. Given the challenges involved in creating PCEs during the COVID-19 pandemic, it may be the case that children who experienced more PCEs did so in spite of countervailing forces, leading to a more impactful relationship between PCEs and mood challenges than would typically emerge outside the context of a global pandemic.

Finally, we cannot establish a causal relationship among the variables due to the cross-sectional design (Savitz & Wellenius, 2023). Prior work suggests that mood challenges and environmental variables such as social support are bi-directional, such that mood challenge symptoms (e.g., withdrawal, reactivity) decrease positive experiences with others, which in turn worsens mood challenge symptoms (Kupferberg et al., 2016; Peterson et al., 2021). The strength of the bidirectional relationship between specific PCEs and specific mood challenge symptoms could be a fruitful direction for future research because it could indicate which PCEs are more likely to remain constant in response to mood challenge onset (e.g., extended family support) compared to others (e.g., peer relations) (Huang et al., 2022).

On a related point, due to the cross-sectional study design, we were not able to measure possible mediators to better understand the processes by which mood challenges were more or less likely. It is important to note that anger control and affect dysregulation may actually lead to future anxiety and depression if not addressed (Sloan et al., 2017). It may be the case that PCEs were relatively less beneficial regarding anxiety and depression because sustained and unaddressed anger control and emotional dysregulation had not previously been identified and targeted, thereby leading to an internalized set of emotion regulation strategies, which are then relatively less amenable to the benefits of PCEs. Understanding these potential processes should be a priority for future work in this area among youth in the child welfare system.

## Conclusion

Adverse experiences can increase the risk for mood challenges, while positive experiences appear to act as protective factors against these challenges. Our study suggests that adverse and positive childhood experiences can have varying effects depending on the mood challenge indicator examined, with modest differences across demographic groupings. Our findings indicate the powerful influence of PCEs as protective factors, particularly surrounding emotional regulation, in the face of high rates of ACEs. These findings support efforts by practitioners to increase PCEs across the full range of children’s social ecologies.

## Supporting information

Supplemental Figure 1. Cumulative Effect of PCE and ACE - Total Sample (Hi-Res)

Supplemental Figure 2. Cumulative Effect of PCE and ACE - Stratified (Hi-Res)

Supplemental Figure 3. Proportion of Youth with ACE and PCE by Age

Supplemental Table 1. Description of Variables

Supplemental Table 2. Results of Main, Sensitivity, and Stratified Analysis

## Data Availability

The source data are not available to the public due to patient information protection requirement.

