## Supplementary figures and images for "The Role of Adverse and Positive Childhood Experiences on Mood Challenges in Youth"

### Supplemental Figure 1. Cumulative Effect of PCE and ACE - Total Sample (Hi-Res)

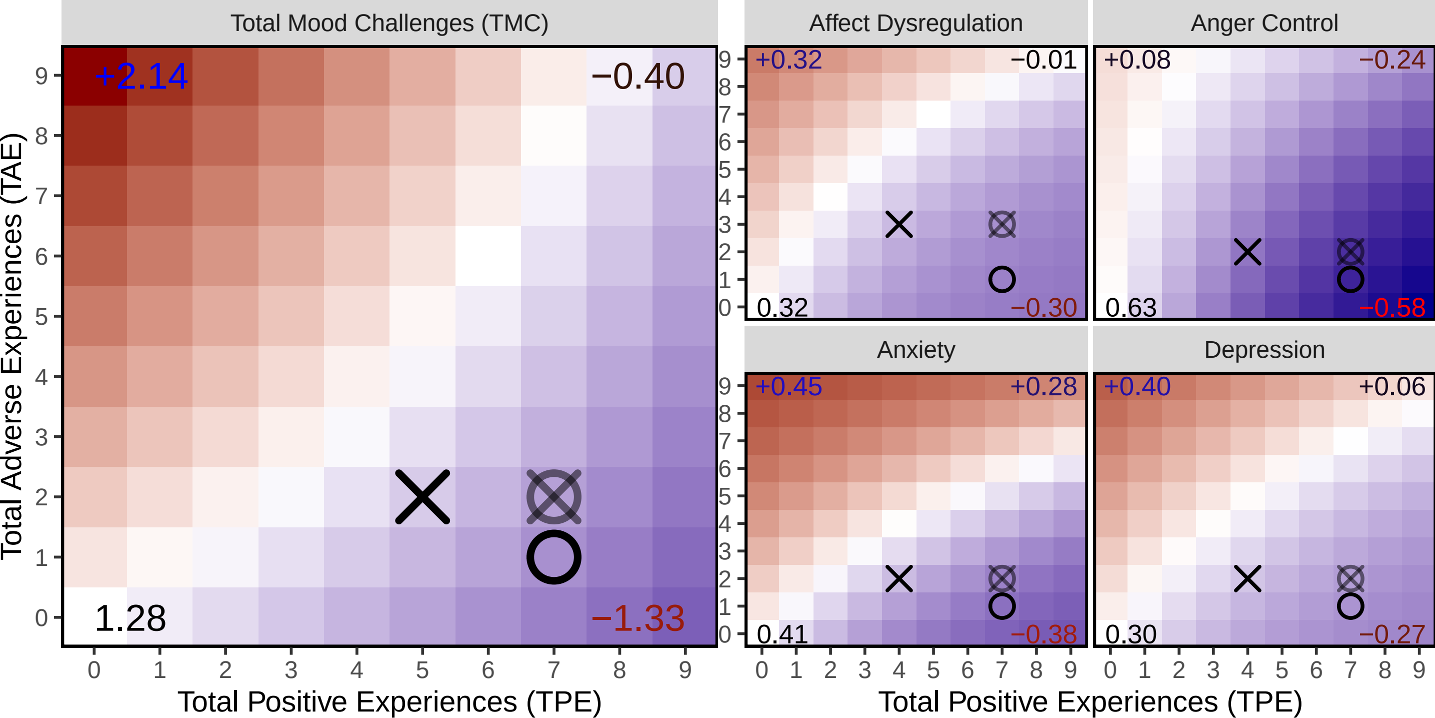

### Supplemental Figure 2. Cumulative Effect of PCE and ACE - Stratified (Hi-Res)

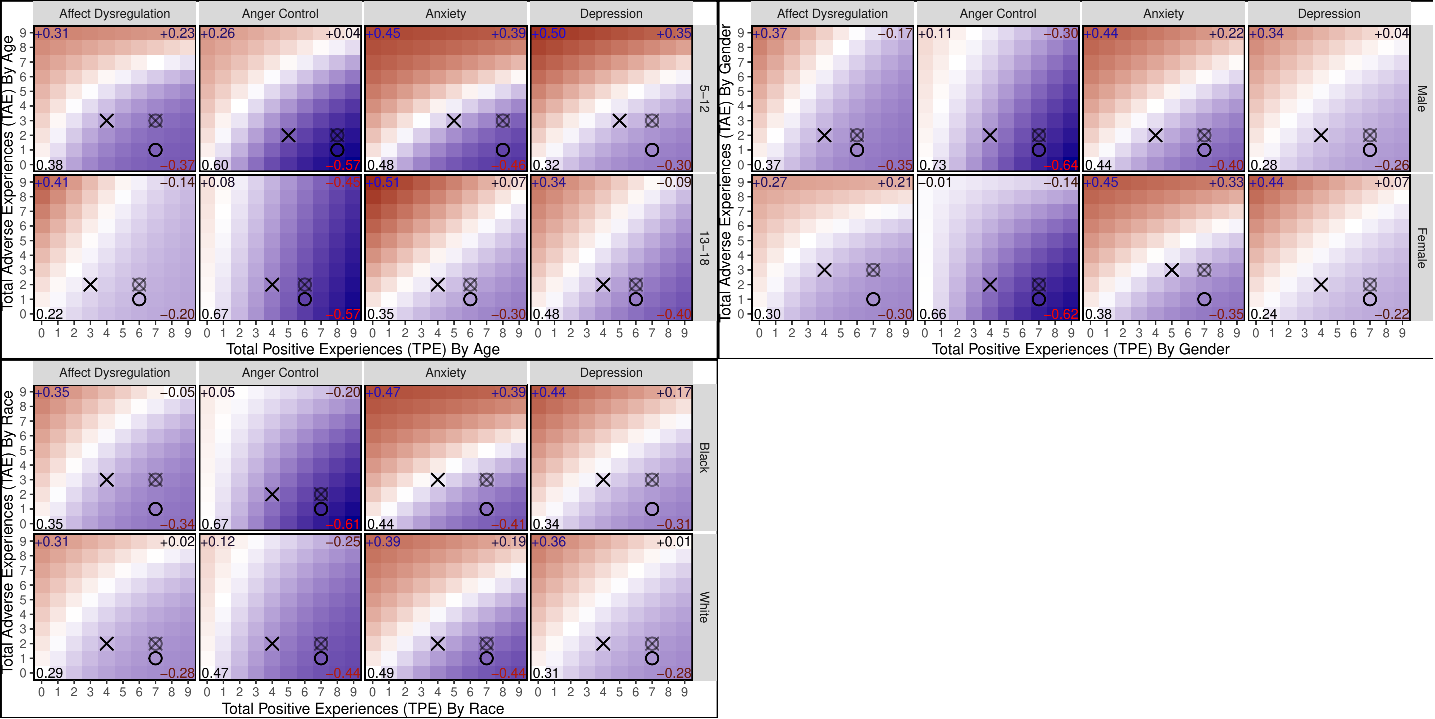

### Supplemental Figure 3. Proportion of Youth with ACE and PCE by Age

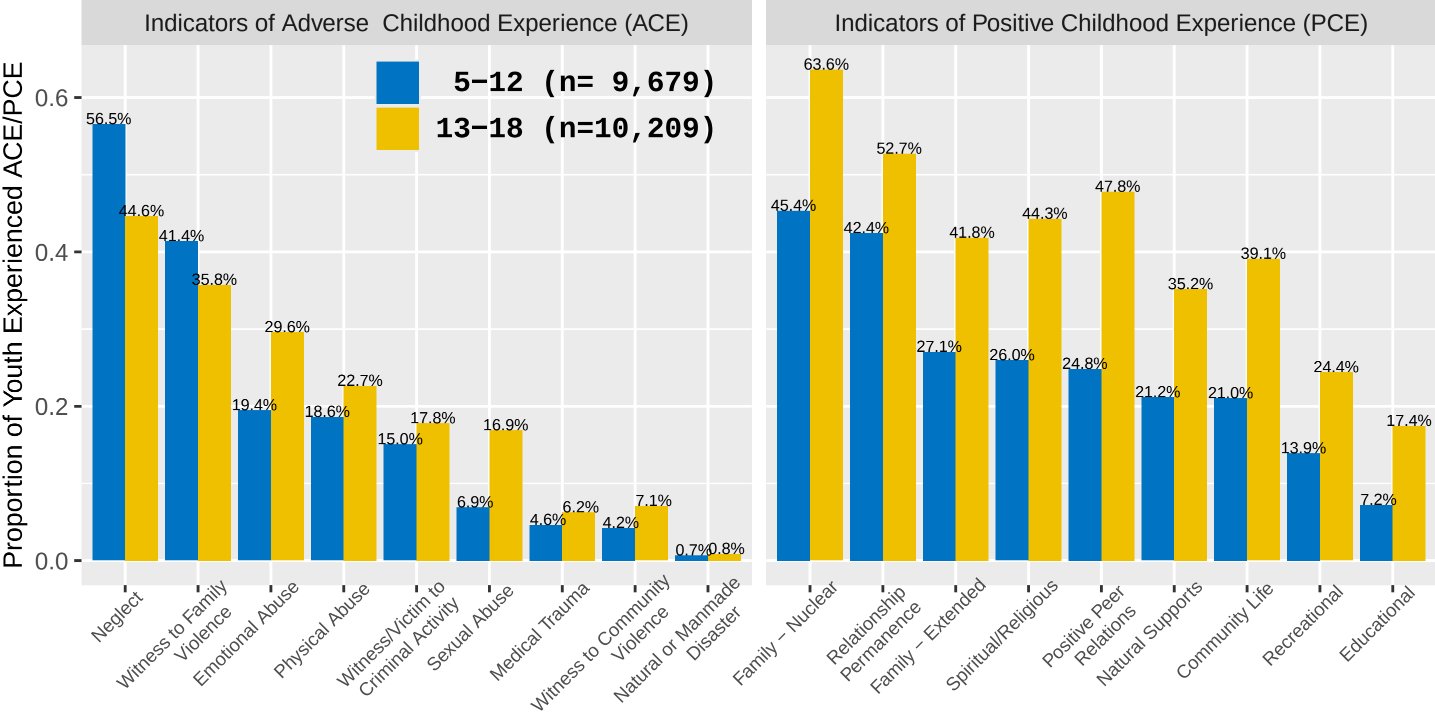
