## Supplemental Methods for "The Role of Adverse and Positive Childhood Experiences on Mood Challenges in Youth"

Details of analysis, interpretation, stratification, and visualization.

### Regression Models

To estimate and compare the adverse effect of ACEs vs the protective effect of PCEs on mood challenge (MC) among the youths, and (2) to explore mutual effect modification (a.k.a., effect on effect, or interaction) between ACEs and PCEs, while controlling for or stratified by demographics (i.e., age, sex, and race), we propose the following default regression model:

L(MC) ~ m + (d) Age + (e) Sex + (f) Race + (a) TAE+ (b) TPE + (c) TAE × TPE

On the right-hand-side, independent variables TAE and TPE are counts of ACEs and PCEs of youth; both are integers ranging from 0 to 9 according to the study definition, and TAE × TPE is simply the product of the two: Age, Sex, and Race are binaries variable defined under “Covariates.” On the left-hand-side, the dependent variable MC represents one of the five outcomes defined under “Mood Challenges and Total Mood Challenge”; “L(.)” is a link function that suits the type the outcome: for any the 4 binary outcomes defined as mood challenge actionable items, L(.) is chosen to be the logit function and the full model is a logistic regression; for the one additional outcome defined as the count of actionable mood challenge items in 0 to 4, L is the identity function and the full model essentially is a linear regression.

### Estimation and Interpretation

The regression coefficients (a) - (f) and (m) were estimated from the study population by R/4.1.2 Statistical Software. In general, (a) and (b) depict the independent effects per additional ACE and PCE on an outcome, respectively; (c) captures the “extra” effect of each ACE in the presence of TPE or of each PCE in the presence of TAE (i.e., mutual effect modification, or effect on effect); therefore, a significantly non-zero (c) means that the effect of each additional ACE or PCE, with independent and modified effect combined, are (a) + (c) TPE and (b) + (c) TAE, respectively, which are not independent unless a youth has not a single ACE or PCE at all; coefficient (d - f) are included as confounding control, which capture the effect of being a post-teen (13-18) vs pre-teen (5-12), being a female vs male, and being a Black vs White, respectively; lastly, (m) represents the grand baseline of the entire study population, and together with (d) Age + (e) Sex + (f) Race, they depict the hypothetical baselines of the 8 demographic groups in the study.

Like the link function, the interpretation of coefficients on TAE or TPE changes from the “odds ratio” of an MC to the “difference in the mean count” of MCs when the number of ACEs or PCEs increases by 1; the meaning of a demographic specific baseline also changes from the “odds” of an MC to the “mean count” of MCs among youth in a demographic group who has zero ACE and PCE.

### Stratified Analysis

Despite controlling for potential demographic confounding via coefficient (d) - (f) and the corresponding variables, the full model assumed constant effects of PCE and ACE across all demographics. To explore age, sex, and race-specific effects, we conducted stratified analysis where a reduced model without the variable defining the strata (e.g., age) is fit twice – once per stratum (e.g., among individuals aged 5-12, then 13-18).

Visualization

A reference grid of 100 cells was created, with both TAE and TPE running from 0 to 9 on the x and y axis, respectively, while Age, Sex, and Race held at reference levels (i.e., 13-18, female, and Black). The grid was plugged into the linear models fitted on the study data to project the outcome (either a probability of an MC, or a count of TMC) at each cell given x number of PCEs and y number of ACEs. Finally, the grid cells were colored by the difference between projected outcome and the baseline outcome given 0 PCEs and 0 ACEs.
