## Supplemental Table 1. Description of Variables for "The Role of Adverse and Positive Childhood Experiences on Mood Challenges in Youth"

Supplemental Table 3.

Description of the outcome and predictor variables defined as CANS (Child and Adolescents Needs and Strengths) indicators rated actionable (i.e., Immediate action/ intensive action required to assist a child).

| **Mood challenge (Behavioral / Emotional)** | Rating for Immediate action/ intensive action required |
|---|---|
| Affect dysregulation | Youth has severe problems regulating affect even with caregiver's support, such as the ease with which someone's mood changes and the intensity of the mood change.<br>• Does the youth have reactions that seem out of proportion to the situation?<br>• Does the youth have extreme or unchecked emotional reactions to situations? |
| Anger control | Youth's temper or anger control problem is dangerous. Youth frequently gets into fights that are often physical. Others likely fear the youth<br>• How does the youth control their emotions?<br>• Does the youth get upset or frustrated easily?<br>• Does the youth overreact if someone criticizes or rejects them?<br>• Does the youth seem to have dramatic mood swings? |
| Anxiety | Clear evidence of f anxiety associated with either anxious mood or significant fearfulness is debilitating, making it virtually it virtually impossible for the youth to function in any life domain<br>• Does the youth have any problems with anxiety or fearfulness?<br>• Is the youth avoiding normal activities out of fear?<br>• Does the youth act frightened or afraid? |
| Depression | Clear evidence of a disabling level of depression that makes it virtually impossible for the youth to function in any life domain. This rating is given to a youth with a severe level of depression. This would include a youth who stays at home or in bed all day due to depression or one whose emotional symptoms prevent any participation in school, friendship groups, or family life. Disabling forms of depressive diagnoses would be rated here<br>• Is the youth concerned about possible depression or chronic low mood and irritability?<br>• Has the youth withdrawn from normal activities?<br>• Does the youth seem lonely or not interested in others? |

| **ACEs (Trauma Experience)** | Rating for Immediate action/ intensive action required |
|---|---|
| Sexual abuse | Youth has experienced severe and repeated sexual abuse, which may have caused physical harm. |
| Physical abuse | Youth has experienced severe and/or repeated physical abuse that causes sufficient physical harm to necessitate hospital or medical treatment |

| | |
|---|---|
| Neglect | Youth has experienced a severe level of neglect, including prolonged absences by adults, without minimal supervision, and failure to provide basic necessities of life on a regular basis |
| Emotional abuse | Youth has experienced severe and repeated emotional abuse over an extended period of time. For instance, the youth is completely ignored by caregivers or threatened/terrorized by others |
| Medical trauma | Youth has experienced life-threatening medical trauma |
| Natural or manmade disaster | Youth has experienced a life-threatening natural or manmade disaster. Youth has been directly exposed to a disaster that caused significant harm or death to a loved one or there is an ongoing impact or life disruption due to the disaster |
| Witness to family violence | Youth has witnessed repeated and severe episodes of family violence. Significant injuries have occurred as a direct result of the violence |
| Witness to community violence | Youth has witnessed or experienced the death of another person in their community as a result of violence; is the direct victim of violence/criminal activity in the community that was life-threatening; or has experienced chronic/ongoing impact as a result of community violence |
| Witness to criminal activity | Youth is a victim of criminal activity that was life-threatening or caused significant physical harm, or the youth witnessed the death of a family friend or loved one |

| **PCEs (Strengths)** | Rating for Immediate action/ intensive action required |
|---|---|
| Relationship permanence | Youth who does not have any stability in relationships. Independent living or adoption must be considered |
| Family-Nuclear | Nuclear family needs significant assistance in developing relationships and communications, OR youth has no identified family |
| Family-Extended | Extended family needs some assistance in developing relationships and/or communications |
| Positive peer relations | There is no evidence of observable interpersonal skills or healthy friendships at this time and/or youth requires significant help to learn to develop interpersonal skills and healthy friendships |
| Educational setting | There is no evidence of the school working to identify or successfully address the youth's needs at this time and/or the school is unable and/or unwilling to work to identify and address the youth's needs and/or there is no school to partner with at this time |
| Recreational | Youth has no recreational opportunities |
| Community life | There is no evidence of an identified community of which youth is a member at this time |
| Spiritual/Religious | There is no evidence of identified spiritual or religious beliefs, nor does the child show any interest in these pursuits at this time |
| Natural supports | Youth has no known natural supports (outside of family and paid caregivers) |
